# Tissue and Peripheral T-cell Repertoire Predicts Immunotherapy Response and Progression-Free Survival in NSCLC Patients

**DOI:** 10.1101/2024.08.01.24311282

**Authors:** Manuel Pino-González, Martín Lázaro-Quintela, Irene Alonso-Álvarez, María Gallardo-Gómez, Laura Juaneda-Magdalena, Alejandro Francisco-Fernández, Silvia Calabuig-Fariñas, Eloisa Jantus-Lewintre, Mónica Martínez-Fernández

## Abstract

Immunotherapy has opened new avenues of treatment for patients with advanced non-small cell lung cancer (NSCLC) without previous hope of survival. Unfortunately, only a small percentage of patients benefit from it, and it is still not well understood which tumor characteristics can be used to predict immunotherapy response. As the key cellular effectors of antitumor immunity, T cells are endowed with specialized T cell receptors (TCRs) to recognize and eliminate cancer cells. Here, we evaluated the potential of TCR repertoire as a predictive biomarker in patients treated with immunotherapy. With this aim, advanced NSCLC patients treated with immunotherapy at first-line were included. After obtaining peripheral blood and tissue samples at baseline, next-generation sequencing targeting TCRβ/γ was performed. Beyond TCR metrics, clonal space of the most frequent clones was determined. We found a positive association between uneven tumor-infiltrating TCRβ repertoire and the immunotherapy response. Moreover, the use of various tumor-infiltrating and circulating TRBV/J genes predicted the immunotherapy response. Our results indicate the importance of evaluating tissue and circulating TCRβ repertoire prior immunotherapy, showing it as a promising immunotherapy response biomarker in NSCLC patients.

## Introduction

According to the latest estimates from the International Agency for Research on Cancer (IARC), lung cancer is the most common and deadliest cancer worldwide [1]. Regarding its histology, it can be classified into non-small cell lung cancer (NSCLC) and small cell lung cancer (SCLC). NSCLC accounts for 85% of cases and is further subdivided into adenocarcinoma, squamous cell carcinoma, and large cell carcinoma [2–4]. Although environmental exposures (such as fuels, arsenic, radon, and air pollution) and medical conditions (as chronic obstructive pulmonary disease, HIV infection, or presence of driver mutations), contribute to its incidence and mortality, smoking is the main risk factor associated with lung cancer [2,3,5].

NSCLC is a highly heterogeneous disease with nonspecific symptoms, leading to a common late diagnosed at advanced stages (III-IV), where surgery is not feasible [2,4,5]. This greatly compromise the patient’s prognosis, which is clearly reflected in the 5-year survival rate, decreasing from 60% in early stages to 4-6% in metastatic settings [2,4,5]. In fact, 90% of cancer deaths are due to metastasis [6].

Nowadays, targeted therapy against many actionable mutations (*EGFR, ALK, RET, BRAF, ROS1, NTRK, MET*, *HER2* and *KRAS*) represent the standard treatment for molecularly defined populations [2,7]. Unfortunately, only 25% of patients carry any of these mutations [2,8]. In 2015, the combination of immunotherapy based on immune checkpoint inhibitors (ICIs) and chemotherapy (CTX) as first-line treatment significantly improved overall survival (OS) in NSCLC patients compared to those treated with CTX alone [2,9,10]. Thus, ICIs has been established as the standard treatment for patients with advanced NSCLC who do not have actionable mutations, either as monotherapy or in combination with CTX, leading to a 5-year survival rate up to 20% [2,9,10]. Unfortunately, only a small percentage of patients (20-40%) benefit from it [2,9,10]. Its high cost, the limited time for therapy selection, and the possibility of developing autoimmune-related side effects (such as inflammatory arthritis, colitis and pneumonitis) underscore the importance of identifying those patients who can truly benefit from the treatment [2,9,10].

Currently, PD1/PD-L1 immunostaining is the only approved predictive biomarker for immunotherapy response, so patients with a tumor proportion score (TPS) ≥ 50% are classified as PD-L1 positive and thus, expected to respond [2,9–11]. However, its reproducibility has been limited by the heterogeneity of results obtained in subsequent studies, largely due to lack of standardization (calculation systems, thresholds, and use of different antibodies) [2,9–11]. Over time, other alternatives such as tumor mutational burden (TMB), tumor-infiltrating lymphocytes (TILs) and other molecular characteristics have been proposed, but none have been able to reliably predict patient response in the real clinical practice [2,9–11].

Although the antitumor response is mediated by different cellular populations, cytotoxic T cells play a key role by recognizing neoantigens and killing cancer cells [12,13]. Neoantigens are recognized by the T cell receptors (TCRs). This transmembrane glycoprotein is composed of an α and a β chain in Tαβ cells (95%) or by a γ and a δ chain in Tγδ cells (5%). Its synthesis occurs through a somatic, stochastic, and imprecise recombination of various non-contiguous homologous gene sets during T cell differentiation in the thymus [11,13–17]. Each chain consists of a variable and a constant region. The variable region of the α and γ chains results from recombination between the V and J genes, while β and δ chains results from recombination between the V, D, and J genes. The result of this V(D)J recombination creates a variable domain that is transcribed and joins one of the constant genes (C), resulting in a functional chain. Each chain contains three hypervariable loops (CDR1, CDR2, and CDR3) in its structure, with CDR3 mediating peptide recognition and thus determining TCR specificity [11,13–17]. CDR3 is encoded by V(D)J genes and undergoes a series of random nucleotide insertions and deletions (N) during recombination at gene junctions (VN(D)NJ). This entire process ensures the creation of a vast array of unique TCRs capable of recognizing a wide range of antigens [11,13–17].

Considering its key role in neoantigen recognition, TCR repertoire analysis provides valuable information about the antitumor response [11,12,14,15,18,19]. Furthermore, advancements in sequencing technologies and bioinformatics tools have enabled sensitive and precise characterization of TCR repertoire in various types of tumors, including NSCLC, making it feasible to study as a potential biomarker for immunotherapy response [11,14,15,18,19]. In recent years, various studies have suggested the potential use of the TCR repertoire as a biomarker for immunotherapy response, both for anti-CTLA4 and anti-PD-L1/PD1 therapies in different types of cancer (such as melanoma, breast and liver), including NSCLC [20–39]. These studies have yielded contradictory results but indicate a potential capacity for the TCR repertoire to be used as an immunotherapy response biomarker. Consequently, the main objective of this study is to evaluate the potential of TCR repertoire analysis as a biomarker for response to immunotherapy in patients with advanced NSCLC.

## Materials and Methods

### Study design and sample collection

This study included twelve patients diagnosed with advanced (IIIA-IVB) NSCLC treated with pembrolizumab either as first-line (N = 11) or second-line treatment (N = 1) at Álvaro Cunqueiro Hospital in Vigo. The study was conducted with appropriate authorization from the Galician Regional Research Ethics Committee (2019/046) following the Helsinki Declaration of 1975 and all patients signed informed consent approving their participation. Patients were classified as responders (R: those showing complete/partial response or stable disease) and non-responders (NR: those with tumor progression or unable to be evaluated due to death) regarding the Response Evaluation Criteria for Solid Tumours (RECIST) [40] at 3 months. To determine both tumor and peripheral TCR repertoire, formalin-fixed paraffin-embedded (FFPE) tissue samples were obtained from all patients prior to immunotherapy and blood samples were drawn from 10 out of 12 patients before the first ICIs dose. Peripheral blood mononuclear cells (PBMCs) were isolated from blood samples by gradient density centrifugation using Ficoll® (Sigma-Aldrich) and cryopreserved until use. DNA was chosen over RNA for analysis due to its superior stability in FFPE samples.

### DNA Extraction

DNA extraction from PBMCs was performed using the QIAamp® DNA Blood Mini Kit (QIAGEN) following the manufacturer’s instructions. DNA from FFPE samples was extracted using the AllPrep® DNA/RNA FFPE Kit (QIAGEN) following the manufacturer’s instructions. DNA quantification was carried out using NanoDropTM 2000c Spectrophotometer (Thermo Fisher Scientific).

### Library preparation and TCR sequencing

A total of 250 ng of genomic DNA was used to prepare the libraries for TCR CDR3β/γ region sequencing following the Oncomine™ TCR Pan-Clonality Assay (Thermo Fisher Scientific). This targeted next-generation sequencing (NGS) assay has been specifically designed to sequence the FR3-J regions of the TCR beta and TCR gamma chains. Libraries were pooled on an Ion 540 chip at 25 pmol/L and sequenced using an Ion GeneStudio S5 Plus Series (Thermo Fisher Scientific).

### Data analysis

After sequencing, data analysis was conducted using Ion Reporter version 5.20.2.0 (Thermo Fisher Scientific). Regarding quality control, off target and unproductive reads were removed. Read classification is shown in *Supp. File S1*. Afterwards, the software reported the VDJ rearrangements and the main repertoire metrics, such as richness, convergence, diversity, and evenness. TCR richness represented the total number of clones, defined as unique TCR beta/gamma nucleotide sequences. TCR convergence was determined as the aggregate frequency of clones which shared a variable gene and CDR3 amino acid sequence. Diversity (Shannon’s diversity) and evenness (normalized Shannon’s diversity) were calculated using the formulas below, in which *p_i_* is the frequency of clone *i* for the sample with *n* unique clones. Evenness describes how evenly distributed is the TCR repertoire. It ranges from 0, meaning the repertoire is unbalanced by a reduced number of predominant clones, to 1, meaning the repertoire is balanced with similar frequencies of all the clones.

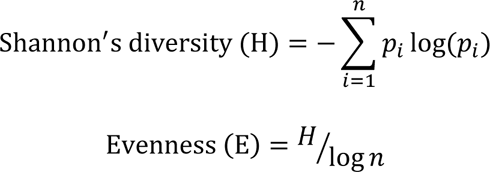

In addition, clones were classified according to their relative abundance in the repertoire, named total clonal space, for each sample. We divided it into top 1% clonal space, top 3% clonal space and top 5% clonal space, so the top 1% clonal space is defined as the aggregate frequencies of the top 1% most frequent clones. All the variables were categorized as “high” or “low” based on their median value.

### Statistical analysis

Statistical analyses were performed using R-Studio version 2023.12.0 and Bioconductor version 3.19 environment. Statistical significance was considered by an overall p-value < 0.05. TCR repertoire variables were correlated with clinical-pathological data (age, sex, tumor histology, PD1/PD-L1 expression, and smoking status), clinical response and survival data (OS and PFS). OS was defined as the time from treatment initiation to death or last follow-up, and PFS was defined as time from treatment initiation to progression or death (whichever is earlier), or last follow-up. Mann-Whitney U test and Student’s t-tests were used to study the relationship between TCR repertoire variables and clinical-pathological data and clinical response. Choice of appropriate test (Mann-Whitney or Student’s t) was based on normality (Shapiro-Wilk test) and variance homogeneity (Levene test). Spearman rank non-parametric test was used for variable correlations. Contingency table analyses when comparing clinical-pathological data with clinical response or clinical-pathological data with TCR repertoire variables were done using Fisher exact test. Kaplan-Meier survival analyses and Log-Rank test were used for time-dependent variables such as OS and PFS. Receiver operating characteristics (ROC) curve analyses were performed to evaluate response prediction, and area under the curve (AUC), sensitivity and specificity values were obtained. The Youden Index method was used to select the best cut-off values [41]. Random Forest Analysis (number of trees = 1000) was used to evaluate the combined predictive signatures performances.

## Results

### Age is associated with immunotherapy response and tumor-infiltrating evenness

This retrospective observational study included a total of twelve patients diagnosed with locally advanced or advanced NSCLC (IIIA-IVB) treated with pembrolizumab at first-line (N = 11) or second-line (N = 1). The most relevant clinico-pathological characteristics are represented in *Table 1*. None of the patients had actionable mutations. Both tissue (N = 12) and PBMCs (N = 10) samples were collected before treatment and were subjected to TCR sequencing. We obtained valid TCR data in all cases (100%).

**Table 1:** Clinico-pathologic characteristics of patients.

| Clinico-pathologic characteristics | Responders (N = 5) | Non-responders (N = 7) | Cohort (N = 12) |
| --- | --- | --- | --- |
| **Age (years)** |  |  |  |
| Median (range) | 74 (71-81) | 60 (43-71) | 70.5 (43-81) |
| **Sex** |  |  |  |
| Male | 4 | 5 | 9 |
| Female | 1 | 2 | 3 |
| **Smoking status** |  |  |  |
| Smoker | 2 | 4 | 6 |
| Former smoker | 3 | 2 | 5 |
| Never smoker | 0 | 1 | 1 |
| **Histology** |  |  |  |
| Squamous | 2 | 1 | 3 |
| Adenocarcinoma | 3 | 6 | 9 |
| **Stage** |  |  |  |
| IIIA | 0 | 1 | 1 |
| IIIC | 0 | 1 | 1 |
| IVA | 4 | 0 | 4 |
| IVB | 1 | 5 | 6 |
| **PD1/PD-L1 staining** |  |  |  |
| Positive | 4 | 6 | 10 |
| Negative | 1 | 1 | 2 |
| **Response type** |  |  |  |
| CP | 0 | 0 | 0 |
| PR | 3 | 0 | 3 |
| SD | 2 | 0 | 2 |
| PD | 0 | 4 | 4 |
| NE | 0 | 3 | 3 |
| **PFS (days)** |  |  |  |
| Median (range) | 328 (116-952) | 72 (3-148) | 100 (4-952) |
| **OS (days)** |  |  |  |
| Median (range) | 405 (126-952) | 113 (4-364) | 225 (4-952) |

ADC: adenocarcinoma, CR: complete response, N: number of patients, NE: non-evaluable. PD: progressive disease, PFS: Progression-free survival, PR: partial response, SCC: squamous cell carcinoma, SD: stable disease, OS: overall survival.

First, we evaluated the effect of immunotherapy response on patient prognosis. As expected, R had statistically significant longer PFS (p = 0.015) than NR (*Fig. 1A*). We found the same trend regarding OS (p = 0.057) (*Supp. File S2A*). Interestingly, we detected an association between response and age, where R were statistically significant older than NR (p = 0.008; R age range = 71-81 years; NR age range = 43-71 years) (*Fig. 1B*). All R were above the median age of the cohort (70.5 years) (*Supp. File S2B*). No other association was detected between clinico-pathological characteristics and response or prognosis.

**Figure 1:**
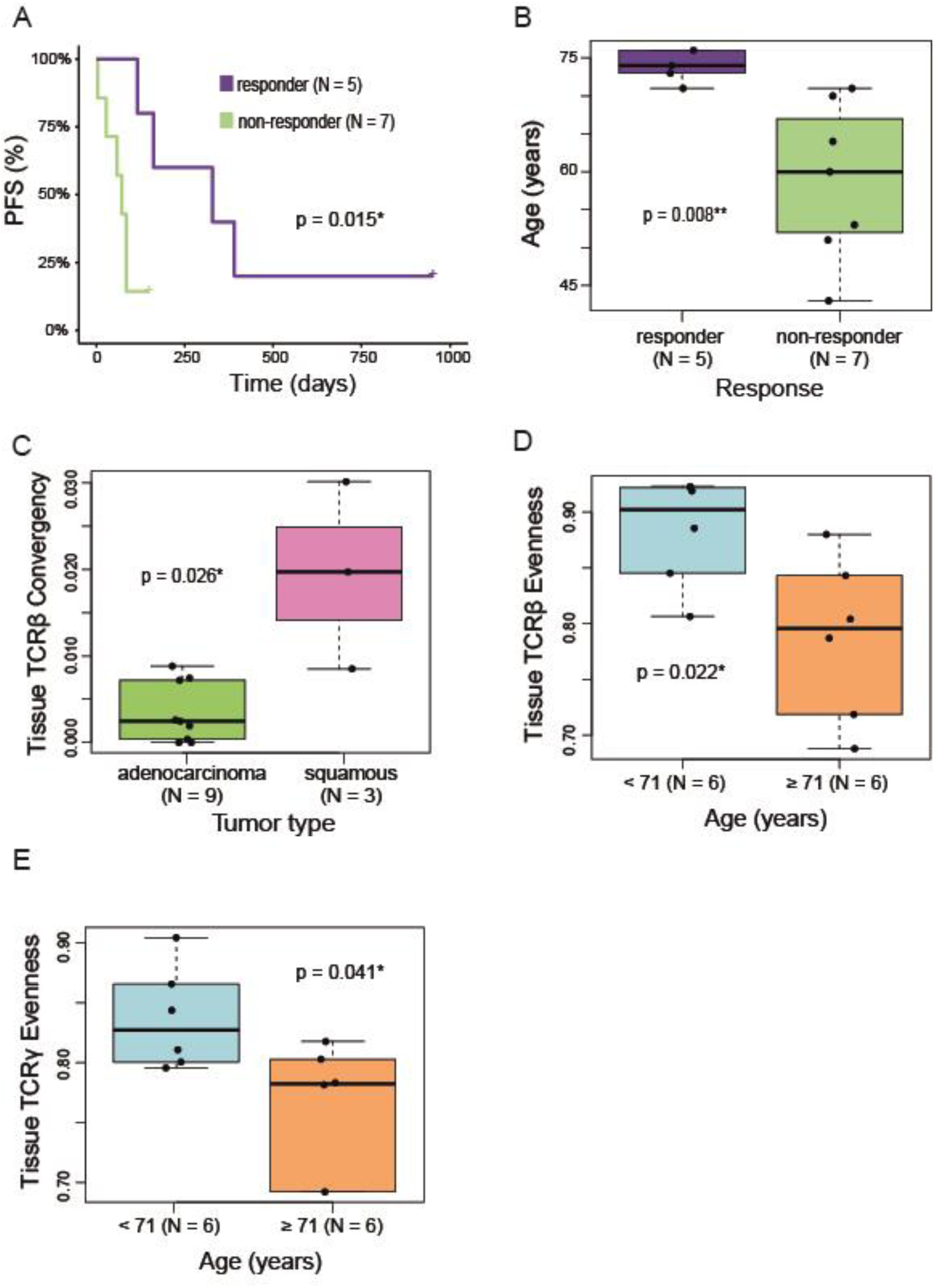
A) Responders (R) have statistically significant longer PFS than non-responders (NR) (p = 0.015). P-value was obtained using Log-Rank test. B) There are an association between response and age, where R are statistically significant older than NR (p = 0.008). P-value was obtained using Student’s t test. C) Squamous have statistically significant higher tumor-infiltrating TCRβ convergence than adenocarcinomas (p = 0.026). P-value was obtained using Mann-Whitney test. D) Younger patients (< 71 years old) have statistically significant higher tumor-infiltrating TCRβ evenness than older patients (≥ 71 years old) (p = 0.022). P-value was obtained using Student’s t test. E) Younger patients have statistically significant higher tumor-infiltrating TCRγ evenness than older patients (p = 0.041). P-value was obtained using Mann-Whitney test. N: number of patients, PFS: progression-free survival, *: statistically significance, **: strongly statistically significance.

Second, we sought for possible associations between the clinico-pathological characteristics and TCR repertoire characterization. We detected that squamous tumors presented statistically significant higher tumor-infiltrating TCRβ convergence (p = 0.026) than adenocarcinomas (*Fig. 1C*). Furthermore, we found an association between age and tumor-infiltrating evenness in both TCRβ and TCRγ, where younger patients (<71 years old) had statistically significant higher tumor-infiltrating evenness than older patients (≥ 71 years old) (p = 0.022 and p = 0.041, respectively) (*Fig. 1D-E*).

### Tumor-infiltrating TCRβ evenness is associated with immunotherapy response and Progression-Free Survival (PFS)

When comparing tumor tissues, we found that R had statistically significant lower tumor-infiltrating TCRβ evenness than NR (p = 0.044) (*Fig. 2A*). Thus, its ability of potentially predicting the response and survival of these patients was analysed. As represented in *Fig. 2B*, tumor-infiltrating TCRβ evenness predicted response with an AUC of 0.86, where a tumor-infiltrating TCRβ evenness lower than 0.795 had a 60% sensitivity and 100% specificity of predicting response. Moreover, patients with low tumor-infiltrating TCRβ evenness (< 0.8441) had statistically significant longer PFS than patients with high (≥ 0.8441) tumor-infiltrating TCRβ evenness (p = 0.013) (*Fig. 2C*). Similarly, we found the same trend regarding circulating TCRβ evenness (p = 0.139) (*Fig. 2D*). Despite they did not reach the statistical significance the same trends were found regarding TCRγ in both tissue and blood, respectively (p = 0.149 and p = 0.114), showing a lower evenness in R patients (*Fig. 2E-F*). It is worth to mention that R patients showed a tendency to a higher tumor-infiltrating and circulating TCRβ top 3% and top 5% clonal space compared with NR patients (*Supp. File S3A-D*). Accordingly, TCRβ evenness and top 3% and 5% clonal space were always strongly negatively correlated both in tissue (R = - 0.81, p = 0.001; R = −0.91, p < 0.001, respectively) and blood (R = −0.99, p < 0.001; R = −0.97, p < 0.001, respectively) (*Supp. File S4*). No other association was detected between clinico-pathological characteristics and TCR repertoire variables.

**Figure 2:**
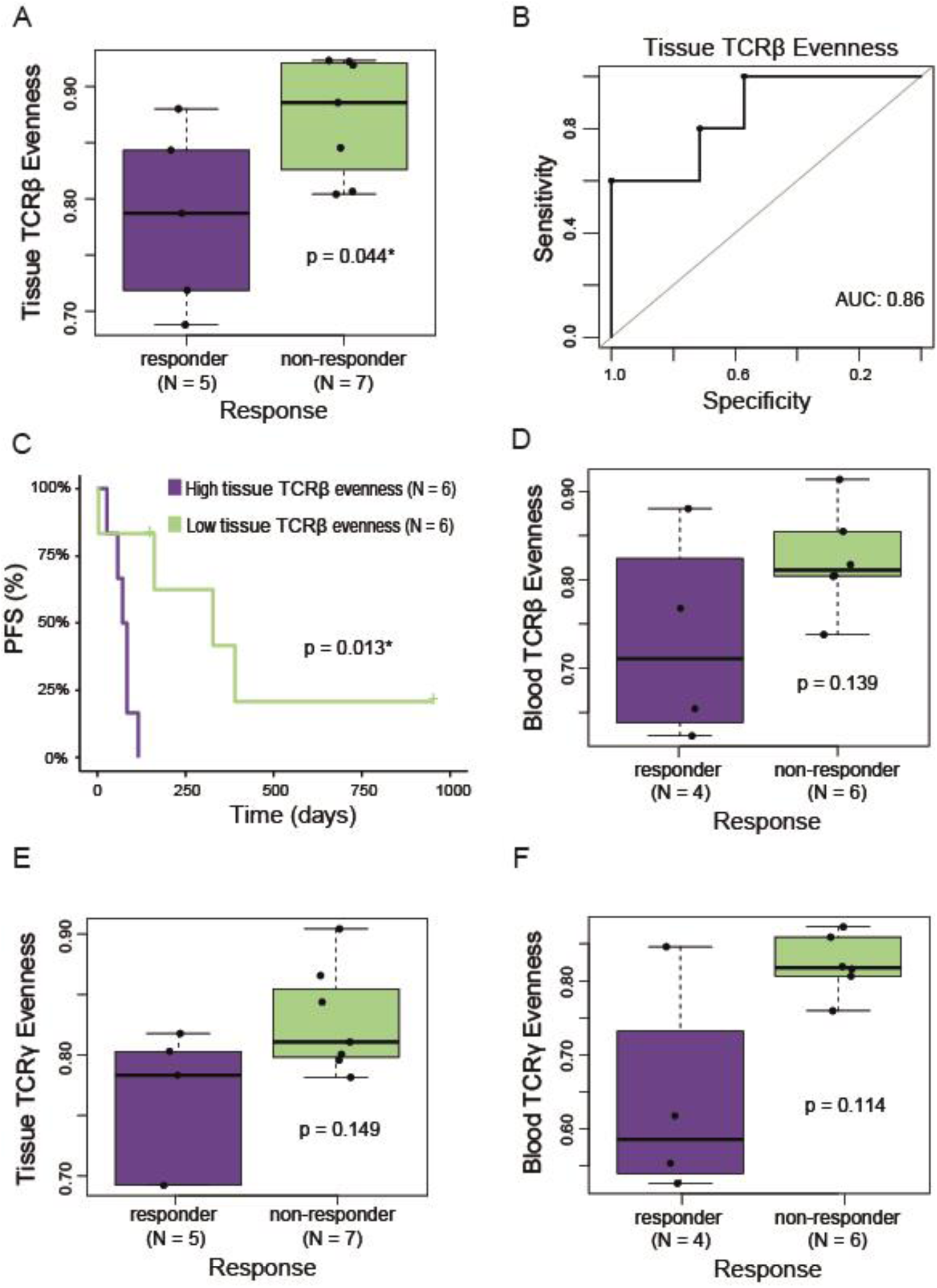
A) Responders (R) have statistically significant lower tumor-infiltrating TCRβ evenness than non-responders (NR) (p = 0.044). P-value was obtained using Student’s t test. B) Tumor-infiltrating TCRβ evenness predicts response with an AUC of 0.86. A lower tumor-infiltrating TCRβ evenness than 0.795 have a 60% sensitivity and 100% specificity predicting response. C) Patients with low tumor-infiltrating TCRβ evenness have statistically significant higher PFS than patients with high tumor-infiltrating TCRβ evenness (p = 0.013). P-value was obtained using Log-Rank test. D) R have a non-statistically significant lower circulating TCRβ evenness than NR (p = 0.139). P-value was obtained using Student’s t test. E) R have a non-statistically significant lower tumor-infiltrating TCRγ evenness than NR (p = 0.149). P-value was obtained using Mann-Whitney test. F) R have a non-statistically significant lower circulating TCRγ evenness than NR (p = 0.114). P-value was obtained using Mann-Whitney test. AUC: Area Under the Curve, N: number of patients, PFS: progression-free survival, *: statistically significance.

### Tumor-infiltrating and circulating TCRβ TRBV and TRBJ genes usage is associated with immunotherapy response and Progression-Free Survival

Next, we sought for any relation between TRBV/TRGV and TRBJ/TRGJ genes frequencies and the immunotherapy response. Regarding TCRγ, there was no association between TRGV and TRGJ genes frequency and clinical response, neither in tissue nor in blood. Interestingly, tumor-infiltrating TCRβ *TRBV6.5*, *TRBV11.3* and *TRBJ2.1* genes frequencies were statistically significant lower in R than in NR (Table 2; p = 0.011, p = 0.048 and p = 0.002, respectively). Moreover, circulating TCRβ *TRBV5.3* gene frequency was statistically significant higher in R than in NR (p = 0.019), while *TRBV27*, *TRBV28*, *TRBJ2.1* and *TRBJ2.6* genes frequencies were statistically significant lower in R than in NR (p = 0.039, p= 0.029, p= 0.017, and p = 0.008, respectively), as shown in *Table 3*.

**Table 2:** Tumor-infiltrating TCRβ TRBV and TRBJ genes frequency in responder (R) compared to non-responders (NR)

| Variable |  | Responder (N = 5) | Non-responder (N = 7) | p-value |
| --- | --- | --- | --- | --- |
| **TRBV6.5** | Median | 0.0082 | 0.0359 | 0.011* |
| **TRBV11.3** | Median | 0.0054 | 0.0077 | 0.048* |
| **TRBJ2.1** | Median | 0.0230 | 0.0581 | 0.002** |

P-value was obtained using Student's t test. N: number of patients, \*: statistically significance, \*\*: strongly statistically significance.

**Table 3:** Circulating TCRβ TRBV and TRBJ genes frequency in responder (R) compared to non-responder (NR). N, number; *, statistically significance; **, very statistically significance.

| Variable |  | Responder (N = 4) | Non-responder (N = 6) | p-value |
| --- | --- | --- | --- | --- |
| **TRBV5.3** | Median | 0.0003 | 0.0001 | 0.019* |
| **TRBV27** | Median | 0.0423 | 0.0897 | 0.039* |
| **TRBV28** | Median | 0.0176 | 0.0633 | 0.029* |
| **TRBJ2.1** | Median | 0.0257 | 0.0662 | 0.017* |
| **TRBJ2.6** | Median | 0.0032 | 0.0092 | 0.008** |

P-value was obtained using Student's t test. N: number of patients, \*: statistically significance, \*\*: strongly statistically significance.

Then, we analysed their ability of potentially predict the PFS and the immunotherapy response in these patients. As represented in *Fig. 3A*, patients with low tumor-infiltrating TCRβ *TRBV11.3* gene frequency presented statistically significant longer PFS than patients with high frequency (p = 0.002). Tumor-infiltrating TCRβ *TRBV11.3* gene frequency predicted response with an AUC of 0.86, where a lower frequency than 0.0057 had an 80% sensitivity and 85.7% specificity predicting response (*Fig. 3B*). In the same line, but not reaching the statistical significance (p = 0.051), patients with low tumor-infiltrating TCRβ *TRBJ2.1* gene frequency had higher PFS than patients with high frequency (*Fig. 3C*). Tumor-infiltrating TCRβ *TRBJ2.1* gene frequency predicted response with an AUC of 0.94, where a lower frequency than 0.0460 had an 100% sensitivity and 85.7% specificity predicting response (*Fig. 3D*). Moreover, patients with low circulating TCRβ *TRBJ2.6* gene frequency had statistically significant higher PFS than patients with high frequency (p = 0.003) (*Fig. 3E*). Circulating TCRβ *TRBJ2.6* gene frequency predicted response with an AUC of 1.00, where a lower frequency than 0.0055 had an 100% sensitivity and 100% specificity predicting response (*Fig. 3F*). Regarding the other genes, despite most of them showed a clear tendency predicting PFS, they did not reach the statistical significance, as represented in *Supp. File S5*. In fact, most of them predicted response with high AUC (0.88-0.96) (*Supp. File S6*).

**Figure 3:**
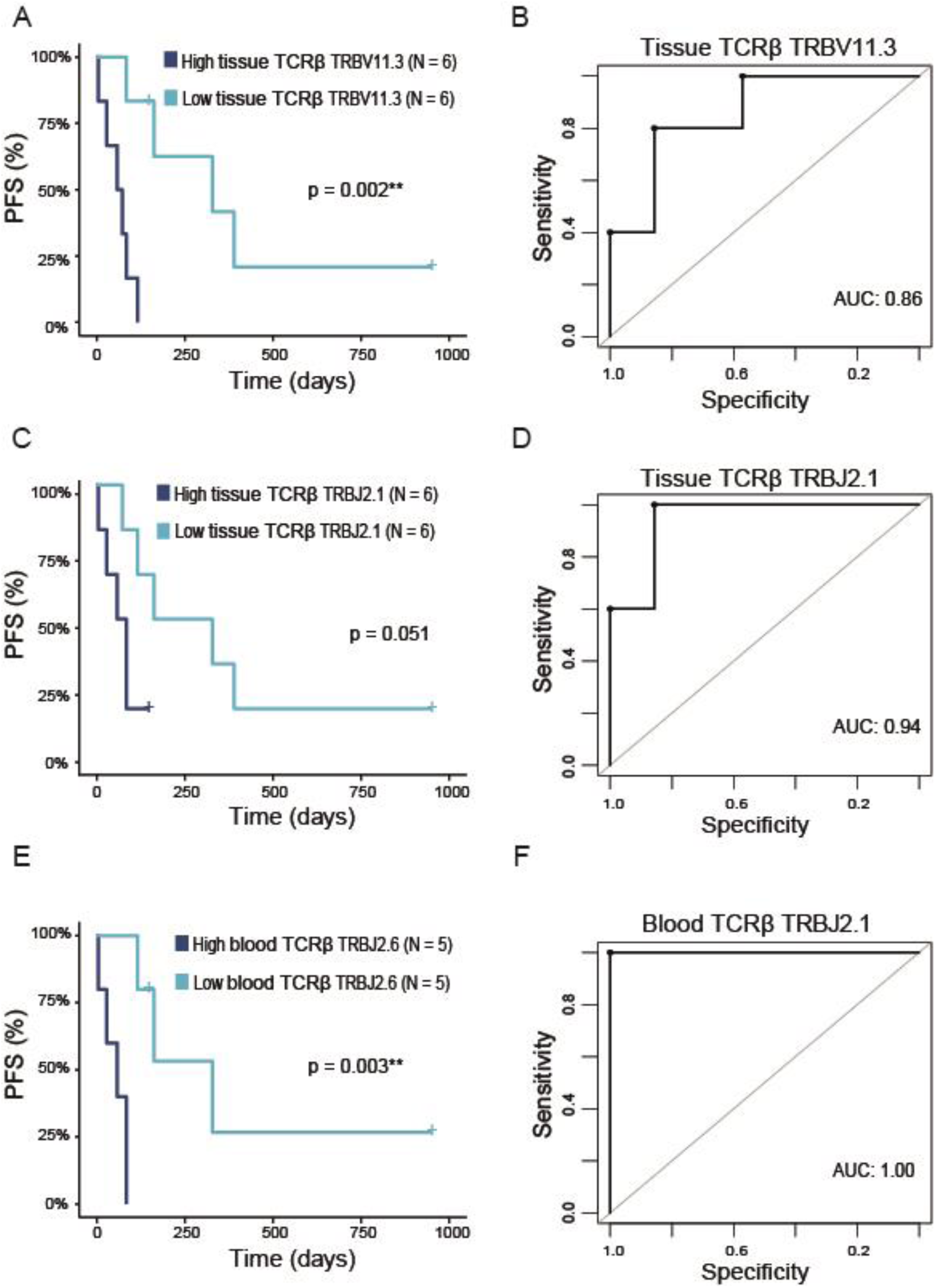
A) Patients with low tumor-infiltrating TCRβ TRBV11.3 gene frequency have statistically significant higher PFS than patients with high tumor-infiltrating TCRβ TRBV11.3 gene frequency (p = 0.002). P-value was obtained using Log-Rank test. B) Tumor-infiltrating TCRβ TRBV11.3 gene frequency predicts response with an AUC of 0.86. A lower tumor-infiltrating TCRβ TRBV11.3 gene frequency than 0.0057 have an 80% sensitivity and 85.7% specificity predicting response. C) Patients with low tumor-infiltrating TCRβ TRBJ2.1 gene frequency have non-statistically significant higher PFS than patients with high tumor-infiltrating TCRβ TRBJ2.1 gene frequency (p = 0.051). P-value was obtained using Log-Rank test. D) Tumor-infiltrating TCRβ TRBJ2.1 gene frequency predicts response with an AUC of 0.94. A lower tumor-infiltrating TCRβ TRBJ2.1 gene frequency than 0.0460 have an 100% sensitivity and 85.7% specificity predicting response. E) Patients with low circulating TCRβ TRBJ2.6 gene frequency have statistically significant higher PFS than patients with high circulating TCRβ TRBJ2.6 gene frequency (p = 0.003). P-value was obtained using Log-Rank test. F) Circulating TCRβ TRBJ2.6 gene frequency predicts response with an AUC of 1.00. A lower circulating TCRβ TRBJ2.6 gene frequency than 0.0055 have an 100% sensitivity and 100% specificity predicting response. AUC: Area Under the Curve, N: number of patients, PFS: progression-free survival, *: statistically significance, **: strongly statistically significance.

### Combined TCRβ signatures to predict response

Although some of these TRBV/TRBJ genes did not reach the statistical significance in predicting PFS, they were still able to accurately predict the clinical response. Based on this observation, we decided to develop two predictive signatures that combine the previously described tumor-infiltrating or circulating TCRβ variables associated with clinical response.

For the tissue predictive signature, we included variables such as evenness, TRBV6.5, TRBV11.3, and TRBJ2.1. By performing Random Forest analyses, this signature was able to predict clinical response with an AUC of 0.83. A positive result from this signature had a sensitivity of 100% and a specificity of 71.4% in predicting response (*Fig. 4A*). In contrast, the predictive signature for blood samples included the variables TRBV5.3, TRBV27, TRBV28, TRBJ2.1, and TRBJ2.6. After conducting Random Forest analyses, this signature demonstrated a predictive ability with an AUC of 0.92. In this case, a positive result had a sensitivity of 75% and a specificity of 100% in predicting response (*Fig. 4B*).

**Figure 4:**
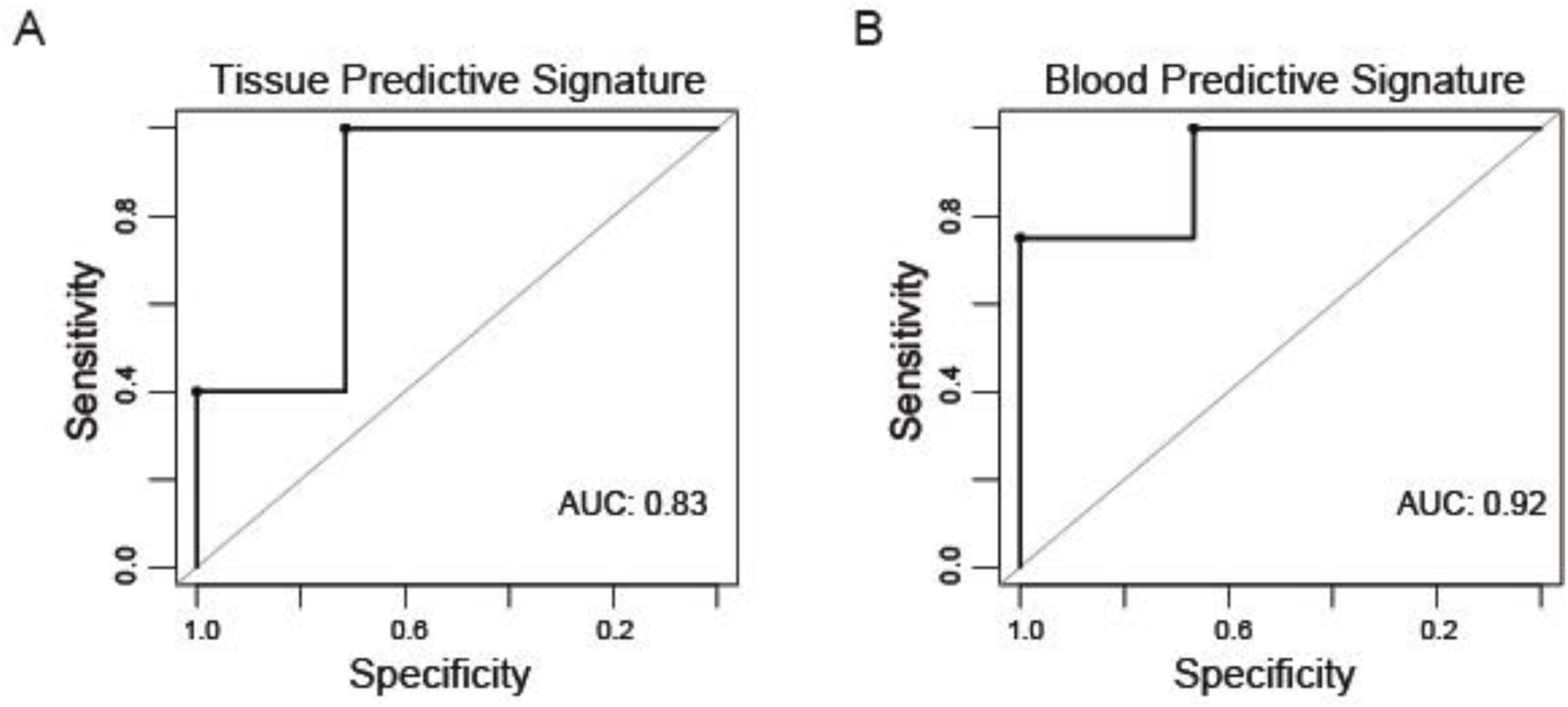
A) A predictive signature including tumor-infiltrating TCRβ variables (evenness, TRBV6.5 gene frequency, TRBV11.3 gene frequency and TRBJ2.1 gene frequency) predicts response with an AUC of 0.83. A positive result has an 100% sensitivity and 71.4% specificity predicting response. B) A predictive signature including circulating TCRβ variables (TRBV5.3 gene frequency, TRBV27 gene frequency, TRBV28 gene frequency, TRBJ2.1 gene frequency and TRBJ2.6 gene frequency) predicts response with an AUC of 0.92. A positive result has an 75% sensitivity and 100% specificity predicting response. AUC: Area Under the Curve.

These findings highlight the potential of using combined TCRβ variables to create effective predictive signatures for both tissue and blood, aiding in the accurate prediction of clinical response.

## Discussion

ICI-based immunotherapy is the standard treatment for advanced NSCLC patients who lack of actionable mutations and cannot undergo surgery. Despite ICIs have significantly improved both life expectancy and quality of life for patients who benefit from it, the response rate is still low (20-40%) [2,9,10]. To date, the lack of a reliable predictive response biomarkers leads to the fact that most of patients are treated with ICIs, highlighting the huge need of identifying those patients who can truly benefit from the treatment and offer alternative treatments to the others [2,9,10]. Recently, the study of the TCR repertoire has emerged as a potential predictive biomarker for immunotherapy response in various types of cancer (such as lung, melanoma, liver or breast) [20–39]. It is noteworthy and important to consider the lack of methodological standardization. Although most of the starting materials, technologies, library preparation approaches, statistical methods and bioinformatics tools used are valid and approved, the combination of all these factors generates variability that greatly complicates the comparison of results [15,18,19,42,43]. Therefore, in this study we employed a reliable and efficient commercial kit for library preparation, along with user-friendly and practical software designed to work with FFPE samples, which are commonly used by clinicians. Additionally, we opted to use DNA instead of RNA due to its superior stability, a particularly important consideration when working with FFPE samples [18,42,43].

To the best of our knowledge this is the first study sequencing the tumor-infiltrating and circulating TCRβ/γ repertoire at baseline in advanced NSCLC patients treated with immunotherapy. Most studies regarding the TCR repertoire have focused solely on Tαβ cells [20–26,29–32,34–39], with very few studying the Tγδ cells, especially in solid tumors [27,28]. Although the mechanism of action of Tγδ cells is still not precisely known and they represent only a small portion of the T cell population, it has been shown that they can play an important role in antitumor response in both systemic and local immunosurveillance [14,17,27,28,33]. For this reason, we decided to include also the γ chain in this work. Moreover, many studies have analysed the TCR repertoire dynamics before and after treatment [20,23,25,26,30–32,34,37,39], providing valuable insights into the mechanisms behind the antitumor response reactivation. However, most advanced NSCLC patients do not survive long enough to start a second-line therapy. Thus, the study of the TCR repertoire as a biomarker for ICIs response should be applicable before the first-line treatment. Therefore, this study focused on the TCR repertoire at baseline.

In this study, we included 12 advanced NSCLC patients treated with ICIs at first and second line. Our results confirmed that R achieved greater PFS, with the same trend observed for OS. Interestingly, we found that R were older than NR. On the contrary, a comprehensive comparative analysis using a cohort of anti-PD1-treated patients with renal cell carcinoma identified no statistically significant difference in PFS or OS with age [44]. However, due the limited numbers of older patients treated with ICIs available for their analyses there is a need for further investigation of this subject in other cancer types, including NSCLC [44]. We also found older patients had a lower TCRβ/γ evenness in tissue but not in blood. In concordance, Erbe *et al.* reported a decrease in tissue TCR evenness with age in patients with adenocarcinoma [44]. Moreover, Dong *et al*. did not find such an association regarding blood either [32]. On the contrary, following a different methodology, Liu et al. found that circulating TCRβ diversity was lower in older patients (>60 years) [26].

Regarding the histological classification and the TCR repertoire, previous studies showed contradictory associations. For instance, some studies described how patients with squamous cell carcinomas had lower tumor-infiltrating TCRβ evenness [24,38], clonal richness [38] or diversity [24] compared to patients with adenocarcinoma. In contrast, and as far as we know for the first time, we observed squamous had higher tumor-infiltrating TCRβ convergence than adenocarcinomas, while others did not find any association between histology and any TCR repertoire metric [31,32]. Although it should be verified in larger cohorts, this disparity might be due to our cohort containing only three squamous cell carcinomas.

Previous studies showed smoking can inhibit T cell proliferation and that smoking duration negatively correlates with TCR evenness [32,38]. While some studies found lower circulating TCRβ clonal richness in smokers [32], others noted higher tumor-infiltrating TCRβ evenness in never-smokers [24]. However, other studies found no association between TCR metrics and smoking [21,29]. Discrepancies may arise from how former smokers are classified. Given that smoking effects persist long-term, we included former smokers with smokers. Due to having only one non-smoker in our cohort, we lacked the statistical power to assess smoking’s impact on TCR repertoire metrics.

Our results showed that R exhibit low tumor-infiltrating TCRβ evenness at baseline, as well as been able of predicting a longer PFS. This could be due to the clonal expansion of the clones mediating the response. Concordant findings were also found in clear cell renal cell carcinoma, where R were observed to have lower tumor-infiltrating TCRβ evenness at baseline [39]. Moreover, Casarrubios *et al*. reported a high tumor-infiltrating TCRβ top 1% clonal space and a low evenness at baseline were involved in a pro-inflammatory tumor microenvironment associated with the complete pathological response of partially advanced stage NSCLC patients treated with neoadjuvant chemo-immunotherapy [34]. Additionally, they described how these same top 1% clones expanded in peripheral blood after treatment, suggesting a possible role of systemic immunosurveillance in preventing relapse. Despite us and Zhang *et al*., found this same trend regarding circulating TCRβ there is no study reporting such association [20,30,32,34,35,39].

In our case, as well as in other studies [26,30,32,34], we did not find any association between TCRβ diversity at baseline and the clinical response. On the contrary, other studies observed patients with greater circulating TCRβ diversity at baseline showed a better response and longer PFS to ICIs + CTX or ICIs alone, highlighting the role of cytotoxic T cells in the antitumor response [25,31,35]. A possible explanation for this disparity could be that in peripheral blood, the abundance of tumor-specific T cell may be diluted by other factors (age, antigen exposure, or immunosuppression), affecting especially the diversity.

After studying the TCRβ gene usage regarding the response and the PFS to immunotherapy, we found some of them (*TRBV5.3, TRBV6.5*, *TRBV11.3, TRBV27*, *TRBV28, TRBJ2.1 and TRBJ2.6*) were associated with the immunotherapy response either regarding tissue or blood. Besides, *TRBV11.3* and *TRBJ2.6* also predicted PFS. Interestingly, R had a higher circulating TCRβ *TRBV5.3* than NR. This could be due some of the neoantigens commonly expressed in the tumor presented high affinity for the sequence encode by this gene. As consequence, it would promote the recognition and eliminations of the tumor cells, mediating the antitumoral response. Similarly, Dong *et al*. reported a circulating TCRβ low *TRBV6.5* and high *TRBV20.1* at baseline showed a longer PFS and OS [32]. Plus*, TRBV20.1* has been reported previously as one of the most used TCRβ V-gene in tumor-infiltrating T cells in several solid tumors [45]. Therefore, it seems the TRBV/J gene usage is also, being able predict the response both in tissue and blood. However, it is not clear which genes are more decisive, if there are ones.

Consequently, we decided to combine all the variables associated with response either in a tumor-infiltrating or circulating predictive signature using a random forest classifier. In both cases, they predicted the response accurately (AUC of 0.83 with 100% sensitivity and 71.4% specificity for tissue, and AUC of 0.92 with 75% sensitivity and 100% specificity for blood), suggesting that a combined signature could be more reliable than a specific variable to predict immunotherapy response.

Regarding TCRγ, we detected the same trend found in TCRβ repertoire without reaching the statistical significance, where R had a lower tumor-infiltrating and circulating TCRγ evenness at baseline than NR. However, we did not find association between TCR variables or V/J genes usage and immunotherapy response. Although a high number of distinct TCRγδ can theoretically be generated, certain rearrangements and chain pairings are substantially over-represented, biasing V(D)J recombination and the selection of functional clones [14]. This leads to various oligoclonal populations predominating in the circulation and in different tissues. Furthermore, there is minimal knowledge reported to date regarding the TCRγ repertoire as an immunotherapy response biomarker [27,28], so further validation will be necessary to verify these findings and draw more definitive conclusions.

It is noteworthy that in our study, we described both tumor-infiltrating and circulating TCRβ gene usage is associated with response and PFS, although different genes are present in each compartment. One possible explanation is that a group of Tαβ cells clones had recognized a series of neoantigens, expanding in peripheral blood and started infiltrating into the tumor, which would explain the low TCRβ evenness found in the tissue. However, some of the clones were remaining in circulation due to the expression of immune checkpoints by the tumor cells. This would change with treatment, allowing all the clones to identify these cancer cells as malignant, thereby promoting their complete infiltration and the elimination of the target. The fact of finding potential biomarkers in blood is particularly important because, although the tumor-infiltrating TCR repertoire can provide more detailed information about the immune response, it can be biased by biopsy due to intratumoral heterogeneity [7,8]. Additionally, finding minimally invasive circulating biomarkers that are easily measurable throughout therapy would facilitate their implementation in daily clinical practice, as the number of recent studies has demonstrated [23,25,26,31,32,35].

## Conclusions

In summary, although much remains to be understood about the antitumor response, this study highlights the potential of TCRβ repertoire analysis as a potential biomarker of response to immunotherapy in NSCLC patients, as we found various tumor-infiltrating and circulating TCRβ variables at baseline able to predict response and progression-free survival. However, the present retrospective observational study is primarily exploratory, aiming to generate hypotheses rather than confirm them definitively. Consequently, further validation in larger cohorts will be necessary to substantiate these findings and for better understanding of the dynamics of the antitumor response.

## Data Availability

The datasets supporting the findings of this study are available from the corresponding authors upon reasonable request.

## Author contributions

MPG performed formal analytics and statistical analysis and wrote the manuscript. MLQ enrolled patients, reviewed the clinical data annotations and wrote the manuscript. IAA performed and optimized DNA isolation and libraries preparation. MGG gave analytics and statistical analysis support. LJM evaluated tissue availability for FFPE sample selection. AFF reviewed the clinical data annotations. SCF and EJL performed TCR sequencing. MMF conceived and designed the study, provided supervision and oversight for the management and execution of this study, acquired financial support for the project, and correct the manuscript. All authors provided critical review, edited the manuscript, and approved the final version.

## Acknowledgements

Authors thank all the enrolled patients and their families. Samples were collected and stored by the Galicia Sur Health Research Institute (IIS Galicia Sur) Biobank (registry B.0000802).

## Funding sources and disclosure of conflicts of interest

This work was supported by the Instituto de Salud Carlos III (ISCIII) and the European Social Fund (“Investing in your future”) (PI21/00348, CP20/00188). MPG is currently supported by a grant for the predoctoral stage (IN606A-2024/017) from the Axencia Galega de Innovación-GAIN, Xunta de Galicia. MGG is currently supported by a grant for the postdoctoral stage (IN606B-2024/014) from the Axencia Galega de Innovación-GAIN, Xunta de Galicia. MMF is currently supported by the Miguel Servet program (CP20/00188) from the Instituto de Salud Carlos III (ISCIII).

The authors declare no competing interests.

## Notes

### Competing Interest Statement

The authors have declared no competing interest.

### Author Declarations

The Galician Regional Research Ethics Committee gave ethical approval for this work (2019/046).

## References

1 Bray F, Laversanne M, Sung H, Ferlay J, Siegel RL, Soerjomataram I, et al. Global cancer statistics 2022: GLOBOCAN estimates of incidence and mortality worldwide for 36 cancers in 185 countries. CA A Cancer J Clinicians. 2024;74(3):229–263.

2 Wang M, Herbst RS, Boshoff C. Toward personalized treatment approaches for non-small-cell lung cancer. Nat Med. 2021;27(8):1345–1356.

3 Leiter A, Veluswamy RR, Wisnivesky JP. The global burden of lung cancer: current status and future trends. Nat Rev Clin Oncol. 2023;20(9):624–639.

4 Lahiri A, Maji A, Potdar PD, Singh N, Parikh P, Bisht B, et al. Lung cancer immunotherapy: progress, pitfalls, and promises. Mol Cancer. 2023;22(1):40.

5 Padinharayil H, Varghese J, John MC, Rajanikant GK, Wilson CM, Al-Yozbaki M, et al. Non-small cell lung carcinoma (NSCLC): Implications on molecular pathology and advances in early diagnostics and therapeutics. Genes & Diseases. 2023;10(3):960–989.

6 Fares J, Fares MY, Khachfe HH, Salhab HA, Fares Y. Molecular principles of metastasis: a hallmark of cancer revisited. Sig Transduct Target Ther. 2020;5(1):28.

7 Tomasik B, Skrzypski M, Bieńkowski M, Dziadziuszko R, Jassem J. Current and future applications of liquid biopsy in non-small-cell lung cancer—a narrative review. Transl Lung Cancer Res. 2023;12(3):594–614.

8 Yang Y, Liu H, Chen Y, Xiao N, Zheng Z, Liu H, et al. Liquid biopsy on the horizon in immunotherapy of non-small cell lung cancer: current status, challenges, and perspectives. Cell Death Dis. 2023;14(3):230.

9 Dugage MR, Albarrán-Artahona V, Laguna JC, Chaput N, Vignot S, Besse B, et al. Biomarkers of response to immunotherapy in early stage non-small cell lung cancer. European Journal of Cancer. 2023;S0959804923000783.

10 Oitabén A, Fonseca P, Villanueva MJ, García-Benito C, López-López A, Garrido-Fernández A, et al. Emerging Blood-Based Biomarkers for Predicting Immunotherapy Response in NSCLC. Cancers. 2022;14(11):2626.

11 Luo H, Wang W, Mai J, Yin R, Cai X, Li Q. The nexus of dynamic T cell states and immune checkpoint blockade therapy in the periphery and tumor microenvironment. Front Immunol. 2023;14:1267918.

12 Jhunjhunwala S, Hammer C, Delamarre L. Antigen presentation in cancer: insights into tumour immunogenicity and immune evasion. Nat Rev Cancer. 2021;21(5):298–312.

13 Sun L, Su Y, Jiao A, Wang X, Zhang B. T cells in health and disease. Sig Transduct Target Ther. 2023;8(1):235.

14 Mensurado S, Blanco-Domínguez R, Silva-Santos B. The emerging roles of γδ T cells in cancer immunotherapy. Nat Rev Clin Oncol. 2023;20(3):178–191.

15 Sanromán ÁF, Joshi K, Au L, Chain B, Turajlic S. TCR sequencing: applications in immuno-oncology research. Immuno-Oncology and Technology. 2023;17:100373.

16 Kabelitz D, Serrano R, Kouakanou L, Peters C, Kalyan S. Cancer immunotherapy with γδ T cells: many paths ahead of us. Cell Mol Immunol. 2020;17(9):925–939.

17 Hu Y, Hu Q, Li Y, Lu L, Xiang Z, Yin Z, et al. γδ T cells: origin and fate, subsets, diseases and immunotherapy. Sig Transduct Target Ther. 2023;8(1):434.

18 Porciello N, Franzese O, D’Ambrosio L, Palermo B, Nisticò P. T-cell repertoire diversity: friend or foe for protective antitumor response? J Exp Clin Cancer Res. 2022;41(1):356.

19 Joshi K, Milighetti M, Chain BM. Application of T cell receptor (TCR) repertoire analysis for the advancement of cancer immunotherapy. Current Opinion in Immunology. 2022;74:1–8.

20 Zhang L. TCR Convergence in Individuals Treated With Immune Checkpoint Inhibition for Cancer. Frontiers in Immunology. 2020;10.

21 Wang Y, Peng L, Zhao M, Xiong Y, Xue J, Li B, et al. Comprehensive analysis of T cell receptor repertoire in patients with KRAS mutant non-small cell lung cancer. Transl Lung Cancer Res. 2022;11(9):1936–1950.

22 Joshi K, De Massy MR, Ismail M, Reading JL, Uddin I, Woolston A, et al. Spatial heterogeneity of the T cell receptor repertoire reflects the mutational landscape in lung cancer. Nat Med. 2019;25(10):1549–1559.

23 Sheng J, Wang H, Liu X, Deng Y, Yu Y, Xu P, et al. Deep Sequencing of T-Cell Receptors for Monitoring Peripheral CD8+ T Cells in Chinese Advanced Non–Small-Cell Lung Cancer Patients Treated With the Anti–PD-L1 Antibody. Front Mol Biosci. 2021;8:679130.

24 Reuben A, Zhang J, Chiou S-H, Gittelman RM, Li J, Lee W-C, et al. Comprehensive T cell repertoire characterization of non-small cell lung cancer. Nat Commun. 2020;11(1):603.

25 Nakahara Y, Matsutani T, Igarashi Y, Matsuo N, Himuro H, Saito H, et al. Clinical significance of peripheral TCR and BCR repertoire diversity in EGFR/ALK wild-type NSCLC treated with anti-PD-1 antibody. Cancer Immunol Immunother. 2021;70(10):2881–2892.

26 Liu Y, Yang Q, Yang J, Cao R, Liang J, Liu Y, et al. Characteristics and prognostic significance of profiling the peripheral blood T-cell receptor repertoire in patients with advanced lung cancer. Int J Cancer. 2019;145(5):1423–1431.

27 Janssen A, Villacorta Hidalgo J, Beringer DX, Van Dooremalen S, Fernando F, Van Diest E, et al. γδ T-cell Receptors Derived from Breast Cancer–Infiltrating T Lymphocytes Mediate Antitumor Reactivity. Cancer Immunology Research. 2020;8(4):530–543.

28 Hunter S, Willcox CR, Davey MS, Kasatskaya SA, Jeffery HC, Chudakov DM, et al. Human liver infiltrating γδ T cells are composed of clonally expanded circulating and tissue-resident populations. Journal of Hepatology. 2018;69(3):654–665.

29 Hu Q, Frank ML, Gao Y, Ji L, Peng M, Chen C, et al. Spatial heterogeneity of T cell repertoire across NSCLC tumors, tumor edges, adjacent and distant lung tissues. OncoImmunology. 2023;12(1):2233399.

30 Han J, Yu R, Duan J, Li J, Zhao W, Feng G, et al. Weighting tumor-specific TCR repertoires as a classifier to stratify the immunotherapy delivery in non–small cell lung cancers. Sci Adv. 2021;7(21):eabd6971.

31 Han J, Duan J, Bai H, Wang Y, Wan R, Wang X, et al. TCR Repertoire Diversity of Peripheral PD-1+CD8+ T Cells Predicts Clinical Outcomes after Immunotherapy in Patients with Non– Small Cell Lung Cancer. Cancer Immunology Research. 2020;8(1):146–154.

32 Dong N, Moreno-Manuel A, Calabuig-Fariñas S, Gallach S, Zhang F, Blasco A, et al. Characterization of Circulating T Cell Receptor Repertoire Provides Information about Clinical Outcome after PD-1 Blockade in Advanced Non-Small Cell Lung Cancer Patients. Cancers. 2021;13(12):2950.

33 Davey MS, Willcox CR, Joyce SP, Ladell K, Kasatskaya SA, McLaren JE, et al. Clonal selection in the human Vδ1 T cell repertoire indicates γδ TCR-dependent adaptive immune surveillance. Nat Commun. 2017;8(1):14760.

34 Casarrubios M, Cruz-Bermúdez A, Nadal E, Insa A, García Campelo M del R, Lázaro M, et al. Pretreatment Tissue TCR Repertoire Evenness Is Associated with Complete Pathologic Response in Patients with NSCLC Receiving Neoadjuvant Chemoimmunotherapy. Clinical Cancer Research. 2021;27(21):5878–5890.

35 Abed A, Beasley AB, Reid AL, Law N, Calapre L, Millward M, et al. Circulating pre-treatment T-cell receptor repertoire as a predictive biomarker in advanced or metastatic non-small-cell lung cancer patients treated with pembrolizumab alone or in combination with chemotherapy. ESMO Open. 2023;8(6):102066.

36 Hogan SA, Courtier A, Cheng PF, Jaberg-Bentele NF, Goldinger SM, Manuel M, et al. Peripheral Blood TCR Repertoire Profiling May Facilitate Patient Stratification for Immunotherapy against Melanoma. Cancer Immunology Research. 2019;7(1):77–85.

37 Robert L, Tsoi J, Wang X, Emerson R, Homet B, Chodon T, et al. CTLA4 Blockade Broadens the Peripheral T-Cell Receptor Repertoire. Clinical Cancer Research. 2014;20(9):2424–2432.

38 Kargl J, Busch SE, Yang GHY, Kim K-H, Hanke ML, Metz HE, et al. Neutrophils dominate the immune cell composition in non-small cell lung cancer. Nat Commun. 2017;8(1):14381.

39 Au L, Hatipoglu E, Robert De Massy M, Litchfield K, Beattie G, Rowan A, et al. Determinants of anti-PD-1 response and resistance in clear cell renal cell carcinoma. Cancer Cell. 2021;39(11):1497–1518.e11.

40 Eisenhauer EA, Therasse P, Bogaerts J, Schwartz LH, Sargent D, Ford R, et al. New response evaluation criteria in solid tumours: Revised RECIST guideline (version 1.1). European Journal of Cancer. 2009;45(2):228–247.

41 Youden WJ. Index for rating diagnostic tests. Cancer. 1950;3(1):32–35.

42 Barennes P, Quiniou V, Shugay M, Egorov ES, Davydov AN, Chudakov DM, et al. Benchmarking of T cell receptor repertoire profiling methods reveals large systematic biases. Nat Biotechnol. 2021;39(2):236–245.

43 Chiffelle J, Genolet R, Perez MA, Coukos G, Zoete V, Harari A. T-cell repertoire analysis and metrics of diversity and clonality. Current Opinion in Biotechnology. 2020;65:284–295.

44 Erbe R, Wang Z, Wu S, Xiu J, Zaidi N, La J, et al. Evaluating the impact of age on immune checkpoint therapy biomarkers. Cell Reports. 2021;36(8):109599.

45 Li B, Li T, Pignon J-C, Wang B, Wang J, Shukla SA, et al. Landscape of tumor-infiltrating T cell repertoire of human cancers. Nat Genet. 2016;48(7):725–732.

