## Supplementary File S1 for "Tissue and Peripheral T-cell Repertoire Predicts Immunotherapy Response and Progression-Free Survival in NSCLC Patients"

Table S1: Description of the type of reads based on their quality. INDEL, Insertion Deletion.

| Thype of read | Description |
| --- | --- |
| **Off-target/low-quality** | Reads that are of low quality or represent the product of an off-target amplification. |
| **Unproductive** | Reads that have uncorrectable sequencing or PCR errors that lead the rearrangement to have out-of-frame variable and joining genes or a premature stop codon. |
| **Rescued productive** | Reads that have an in-frame variable and joining gene, and no stop codons after INDEL error correction. |
| **Productive** | Reads that have an in-frame variable and joining gene, and no stop codons. |
