## Supplementary figures and images for "Tissue and Peripheral T-cell Repertoire Predicts Immunotherapy Response and Progression-Free Survival in NSCLC Patients"

### Supplementary File S2

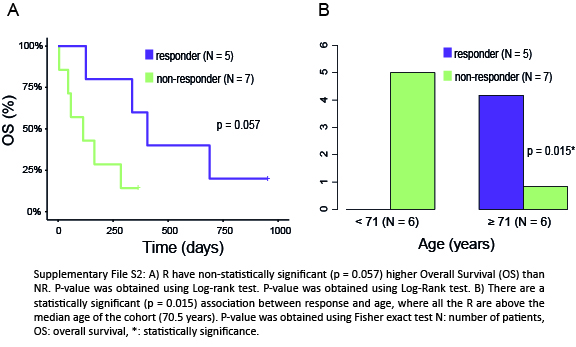

### Supplementary File S3

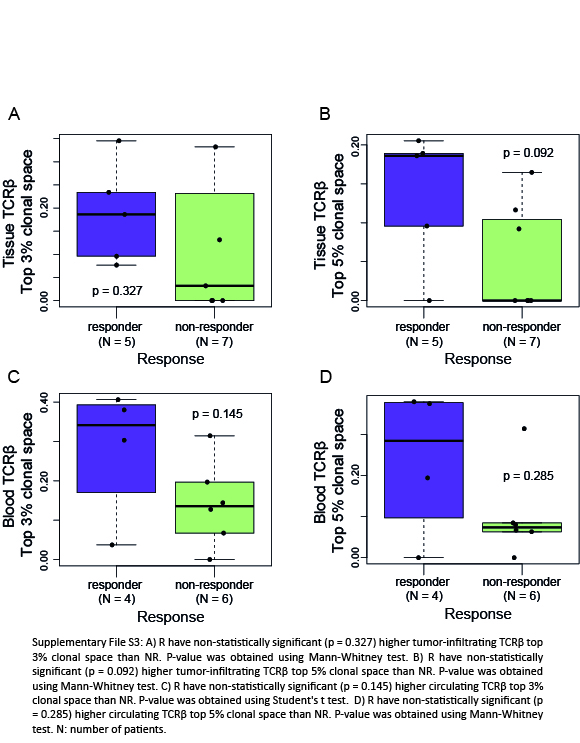

### Supplementary File S4

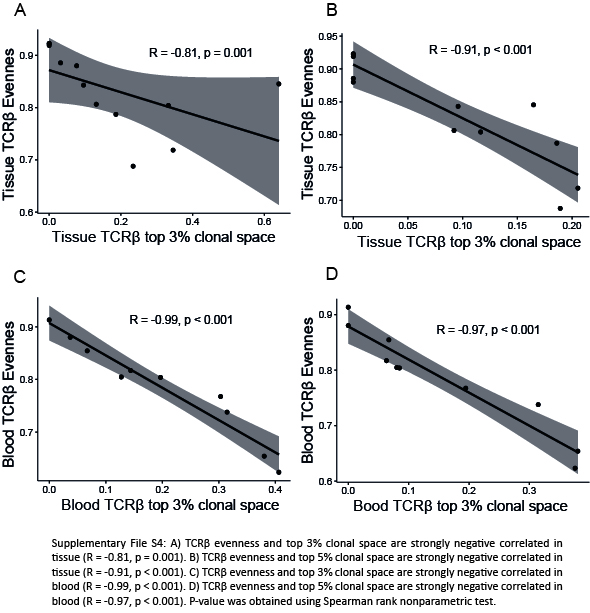

### Supplementary File S5

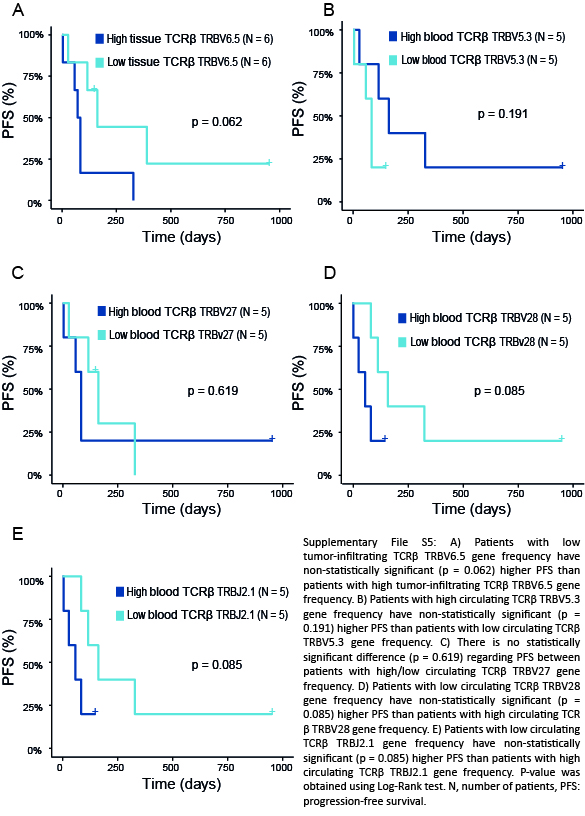

### Supplementary File S6

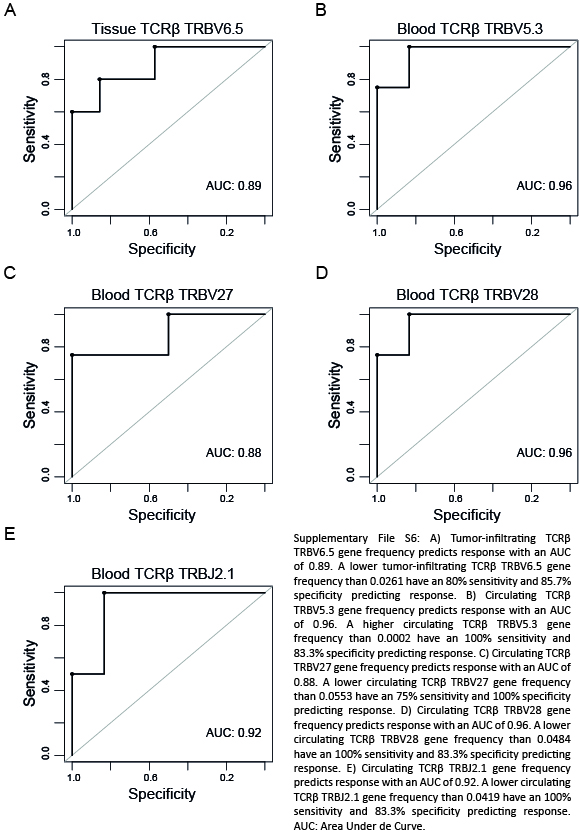
